# Multidisciplinary large language model agent teams for precision oncology enhance complex gynecologic oncology decision support

**DOI:** 10.1101/2025.10.30.25339199

**Authors:** Warisijiang Kuerbanjiang, Xinyu Wang, Yiershatijiang Jiamaliding, Musitapa Maimaitiaili, Yuexiong Yi

## Abstract

Large language models can help with clinical decision-making tasks. Complex oncology cases are best managed through multidisciplinary tumor boards but are difficult to do so due to their expense. The MDAT framework is proposed to mimic tumor board-style collaboration. Some LLMs are prompted to act like experts. They first analyze the prompt from their respective perspectives. Then the decision-making takes place to vote, agree, deal with discord and unify. We evaluated this framework on 3 LLMs: ChatGPT-4o, DeepSeek-R1 and Llama-4 (1,056 clinical questions; 182 cases). It focused on staging and selecting the treatment, managing the complications and following up. Fixed-size MDATs, across all baselines and configurations of MDAT, outperformed six different strong prompting baseline algorithms across all models. DeepSeek-R1 benefits from a five-agent MDAT that performs best (98.26/100). The fixed-size MDATs showed the greatest gains in higher-complexity questions, demonstrating robustness where accurate, multidisciplinary reasoning is most needed. As our MDT-inspired agent enhances LLM accuracy for oncology decision-making, it provides a pragmatic approach to integrating AI into complex cancer management.

**Author summary:** Experts, such as surgeon, oncologist and radiologist get together to decide on the best treatment for a cancer patient but it is rather costly and time consuming. Large language models exhibit excellent performance when it comes to answering questions, particularly in medical contexts. We wanted to discover if large language models can work collaboratively in the same manner. We designed a framework named MDAT, standing for Multidisciplinary Agent Team. Rather than asking a question to one generic AI, our system builds a team of AIs “experts”. Each agent considers a patient’s case from its perspective before all of them work together in a well-designed system to provide the expert recommendation. Through multidisciplinary team (MDT) gynecologic cancer scenarios, we examined this team-based artificial intelligence on a custom-built dataset of more than 1000 questions. According to our findings, this method always provides more reliable and accurate answers than decision-making methods, especially for the toughest cases. According to our findings, we can create safer and more effective ways of helping doctors make crucial decisions for cancer patients by making AI act like a real medical team.

## Introduction

Cervical, ovarian, and endometrial cancers account for a large share of cancer incidence and mortality among women. As a result, gynecologic cancers pose a significant global threat to women’s health[1]. Cervical cancer has a particularly strong impact in low- and middle-income settings. It remains the fourth most common cancer in women worldwide[2]. Beyond epidemiology, the clinical reality is also complex. Treatment decisions often require balancing oncologic control with quality of life, fertility preservation, and pregnancy-related considerations. These factors create a challenging environment. In this setting, evidence must be interpreted within the context of each individual patient and viewed from multiple perspectives[3].

A multidisciplinary team (MDT) for complex tumors has become the accepted standard for complex oncologic decision-making. In gynecologic oncology, optimal care commonly requires the coordinated judgment of surgeons[4], medical oncologists, radiation oncologists, pathologists, radiologists[5], genetic counselors, reproductive medicine specialists[6], and, when pregnancy is involved, obstetricians[7]. However, the expanding volume of guidelines, biomarker driven indications, drug interactions, toxicity profiles, and clinical trial eligibility criteria places sustained pressure on time and attention[8].

Large language models (LLMs) already cover a wide range of medical knowledge[9]. They also display solid proficiency on straightforward diagnostic tasks[10], yet their performance, however, falls as cases become more complex[11]. Accuracy drops further when the supporting evidence is diverse[12]. Single-model systems, even when coupled to retrieval, often struggle with cross disciplinary evidence synthesis, safety constraints, and the need for explicit rationales that clinicians can audit[13]. Multi-agent approaches designed to emulate clinical team deliberation have emerged as a promising direction. In rare disease diagnosis, for example, a multi agent conversation framework outperformed single agent baselines, suggesting that role specialization and peer interrogation may confer advantages over monolithic prompting[14]. However, rare disease triage is not equivalent to oncology decision support, where guideline concordance, molecular actionability, trial eligibility, and contraindication checks must be weighed simultaneously and transparently. The gap between textbook knowledge and practical clinical application in time-pressured settings remains wide[15]. To address this gap, we developed and implemented a Multi-Disciplinary Agent Team (MDAT) framework designed to simulate the collaborative reasoning and evidence appraisal characteristic of a multidisciplinary team[16], which we applied to this study. Our approach uses the shared intelligence from multiple AI agents using different prompting strategies that simulate the different skills and views present in the clinical MDT meeting.

To evaluate its feasibility and safety, the framework was systematically tested under multiple configurations, including comparative analyses of representative LLM and diverse prompting strategies[17], and was applied to a large-scale dataset of consecutive gynecologic oncology cases[18]. The MDAT framework may achieve higher Multidisciplinary team grade concordance and more complete and auditable rationales than single agent baselines while maintaining low rates of contraindication violations. The schematic diagram is shown in Fig 1

**Fig 1.**
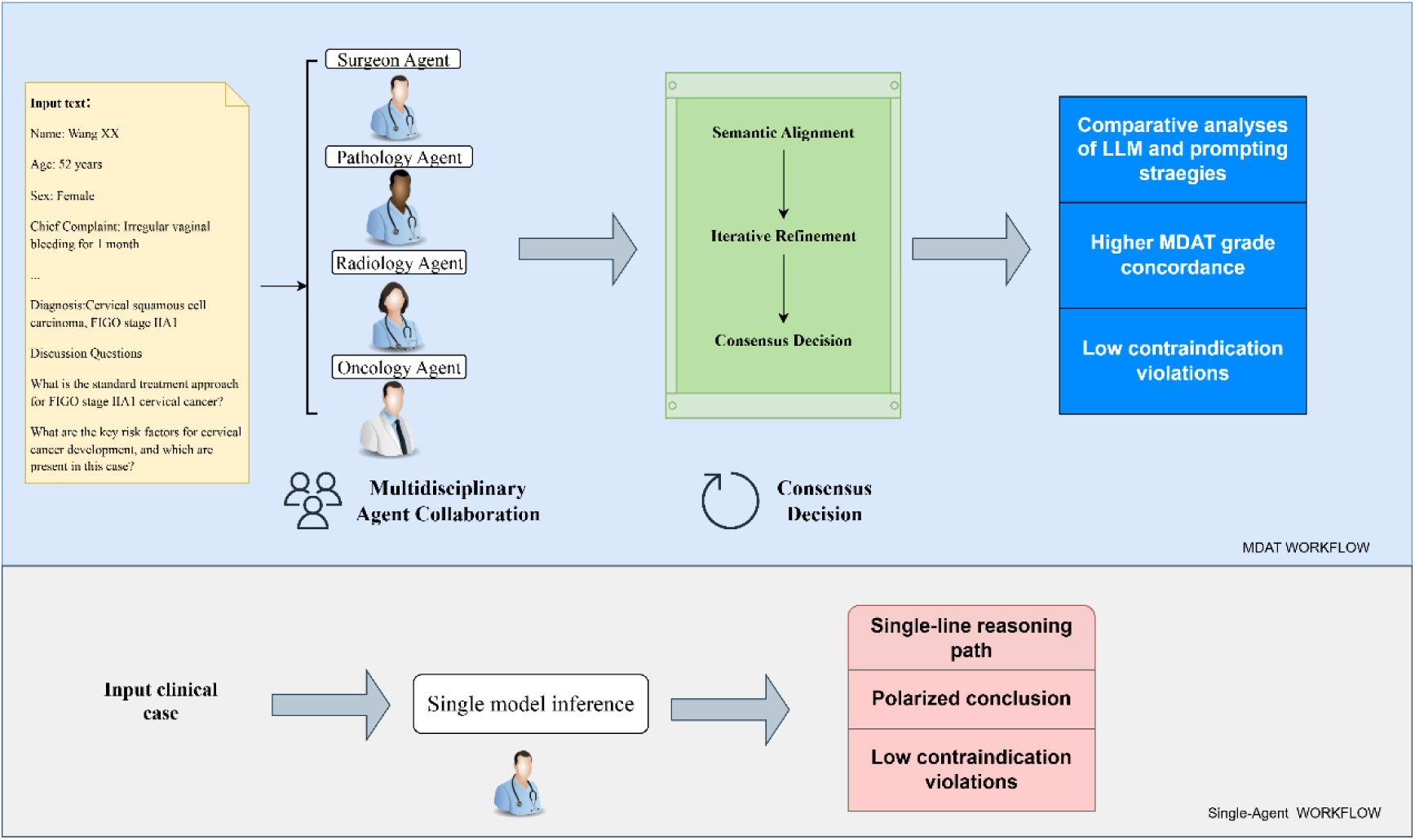
The mechanistic framework of the multidisciplinary agent team (MDAT). Schematic comparing a multi-agent MDT workflow with a single-agent workflow for clinical decision support.

## Result

### MDAT framework implementation

The MDAT succeeds in dynamic role assignment. An innate ability to identify successful collaborative structures is suggested by the experimentally determined ideal team size of 5-7 agents, which the models also autonomously agreed upon. The fact that autonomous agents chose about 5-6 specialists in all three models is an important finding with interesting clinical implications. The selection of a range of team sizes, narrowly clustered around 5.63 to 5.99 agents, by wise agents such as ChatGPT-4o, DeepSeek-R1, and Llama-4.1 agents, who were not directly prompted on what the optimal configuration of agents would be to use in MDT, suggests an understanding of MDT that is more sophisticated than those of the AGENTS providing the prompts. This agent-based selection behavior emulates reality, as gynecologic oncology MDTs typically comprise 5-8 core specialists in clinical practice. A full answer of MDAT is provided in S1 Note.

To make the MDAT system easy for doctors and patients to use, we have also developed a user-friendly graphical interface shown in (Figs S1-3).

### Dataset characteristics and difficulty validation

Our study incorporated 182 gynecologic oncology cases designed to evaluate MDAT performance across multiple clinical reasoning dimensions. The dataset comprised 100 AI-generated cases, developed according to established clinical guidelines and subsequently validated by three medical experts (Fig 2a). This was complemented by 82 cases derived from published case reports that underwent a rigorous expert curation and selection process (Fig 2b). For AI-generated cases, the ICC is 0.71, and for case reports, the ICC is 0.68. Experts show high consistency.

**Fig 2.**
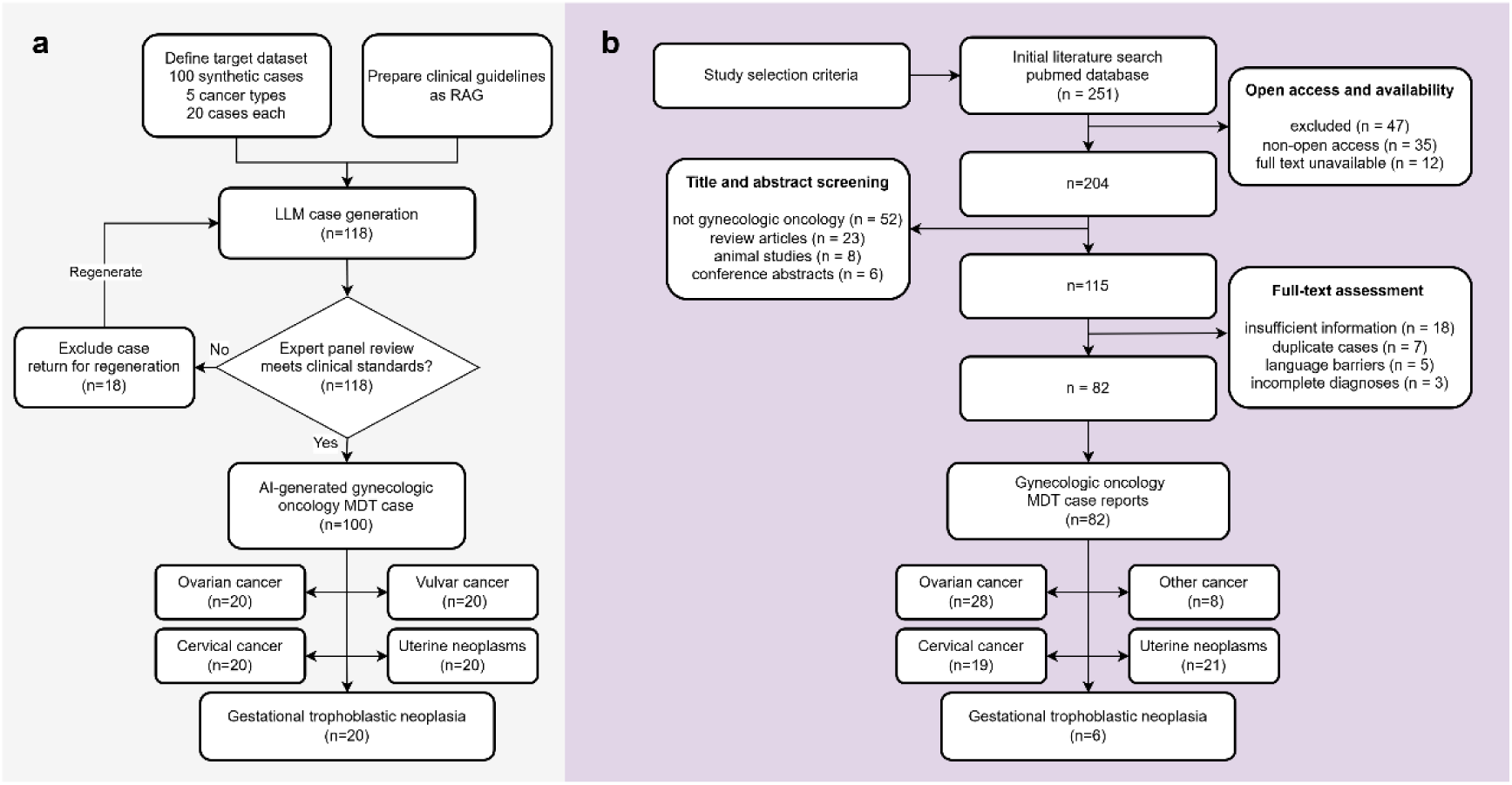
This figure illustrates the dual-source case selection and curation process for the dataset. **a** shows the workflow of AI-generated case (AIGC) group. Dataset of 100 synthetic cases (20 cases each across 5 cancer types) generated with clinical guidelines prepared as RAG, which are distributed among ovarian cancer (n = 20), cervical cancer (n = 20), vulvar cancer (n = 20), uterine neoplasms (n = 20), and gestational trophoblastic neoplasia (n = 20). **b** depicts the case report group workflow, involving systematic literature search from PubMed database (n=251), sequential screening and exclusion processes, and full-text assessment, resulting in 82 gynecologic oncology MDT case reports distributed across ovarian cancer (n=28), cervical cancer (n=19), other cancer (n=8), uterine neoplasms (n=21), and gestational trophoblastic neoplasia (n=6).

The difficulty scores showed distinct distribution patterns between the AIGC and the case report (Fig 3). In 1,056 questions yielded from 182 cases, while the difference in median values is statistically significant, it remains quite small, and the interquartile ranges still overlap in a rather substantial fashion, meaning that effect sizes are relatively modest. Comparison on this dataset is legitimate, as each single model and prompt went through the same questions, meaning it was fair, and all the models are completely compared on the same footing. The difficulty rating scores of the questions show high consistency between two experts with an ICC of 0.61.

**Fig 3.**
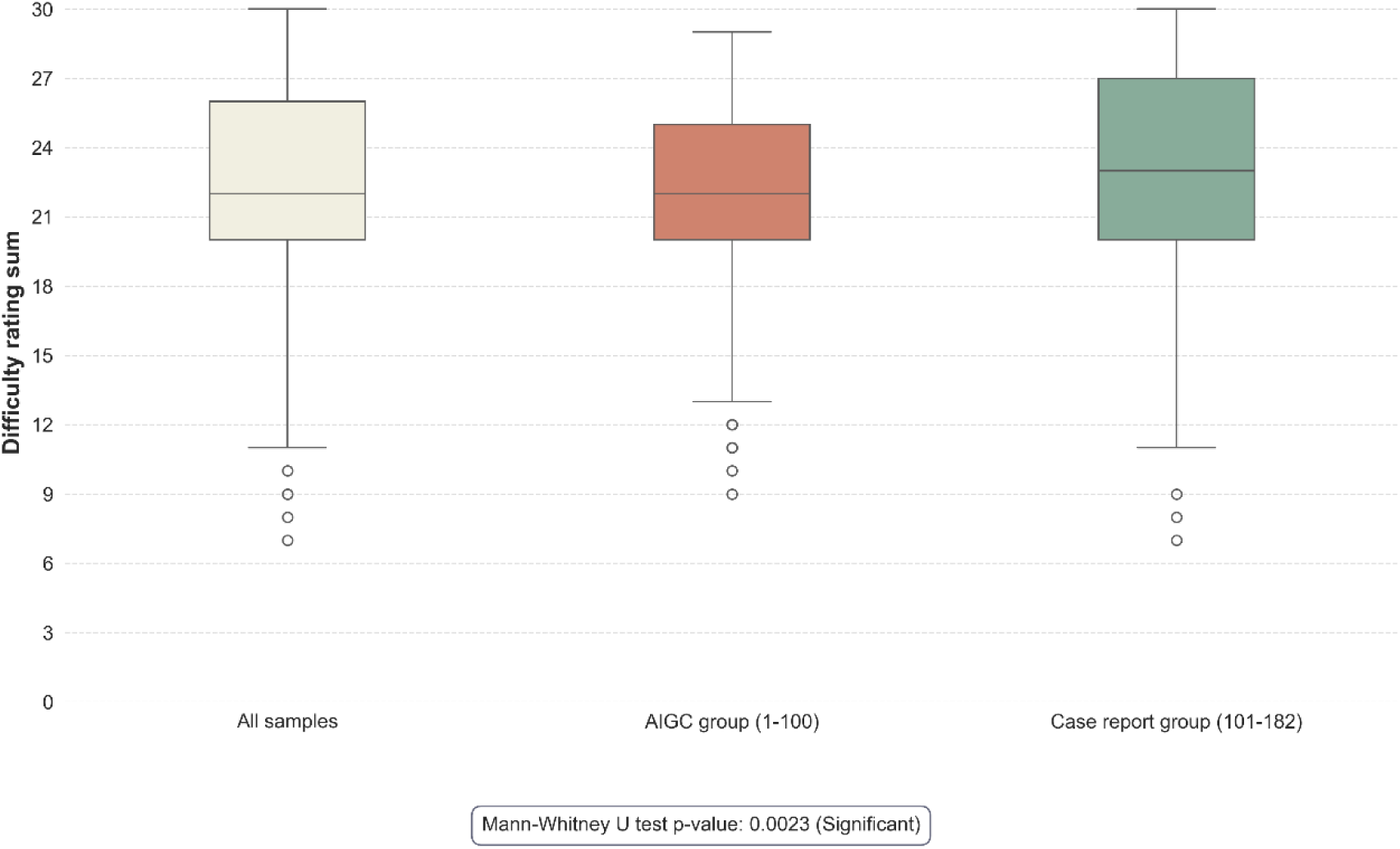
Distribution of Individual Question Difficulty Scores. Comparing the difficulty scores for the entire sample set (“All Samples”), the AI-Generated Content Group ("AIGC Group," cases 1-100), and the human-written “Case Report Group” (cases 101-182). The central line indicates the median score, the box represents the interquartile range (IQR), and the whiskers show the data range, with outliers plotted as individual points. AIGC Artificial Intelligence Generated Content.

### Automated evaluation system validation

Our corpus of 34,848 question–answer pairs was created with 1,056 questions posed to 3 LLMs under 11 prompting strategies, yielding 33 model–prompt pairings. All outputs were rated by an automated evaluation system with a 5-point, 6 dimension Likert-based rubric for accuracy, omissions, potential harm, bias, clarity, and illusion. In order to create a strong reference standard, a subset of 100 replies was independently graded by three clinicians using a harmonized grading scheme resulting in excellent inter-expert reliability (r=0.875; Table 1). Automated evaluation demonstrated high internal consistency (ρ=0.937) and strong agreement with the expert consensus (human–computer agreement r=0.894), supporting its ability to replicate expert-level judgments. Response length did not meaningfully influence quality ratings, as the correlation between token count and automated evaluation scores was negligible (r = −0.0226), indicating that the evaluation effectively mitigates length bias.

**Table 1:** Rubric for Manual Evaluation of Question Difficulty.

| Assessment Dimension | Content Description | Scoring Scale (1-5) |
| --- | --- | --- |
| Diagnostic Complexity | Evaluate the difficulty of diagnosis based on symptoms, differentials, and imaging/pathology. | 1: Very Low Complexity (e.g., textbook case) / 5: Very High Complexity (e.g., atypical presentation) |
| Staging<br>Assessment<br>Difficulty | Assess the complexity of tumor staging using radiological, surgical, and molecular data. | 1: Straightforward (e.g., single definitive finding) / 5: Highly Complex (e.g., integration of multiple data sources) |
| Treatment Decision<br>Complexity | Evaluate the complexity of treatment planning, including surgical and adjuvant therapy choices. | 1: Follows a single, clear guideline / 5: Highly personalized, no clear standard-of-care |
| Multidisciplinary<br>Collaboration | Assess the number of specialties required and the complexity of their coordination. | 1: Managed by a single specialty / 5: Requires complex coordination among many (4+) specialties |
| Prognosis<br>Assessment<br>Challenge | Evaluate the difficulty of prognostic evaluation, including survival and recurrence risk. | 1: Well-established and predictable / 5: Highly uncertain with complex trade-offs |
| Clinical Decision<br>Risk | Assess the risk level of clinical decision-making, including treatment and diagnostic risks. | 1: Minimal risk / 5: High risk of significant adverse outcomes |

### Comparative performance analysis across prompting strategies

Pairwise comparisons across different MDAT configurations revealed significant differences in performance among the three models (Fig 4a). A consistent pattern was observed across all three models, particularly in DeepSeekR1, where the fixed-agent groups exhibited almost non-significant p-values. These teams with 3–9 agents formed a high-performance plateau, indicating that performance remained stable across this range of team sizes. The P value of all pairs is presented in S1 Table.

**Fig 4.**
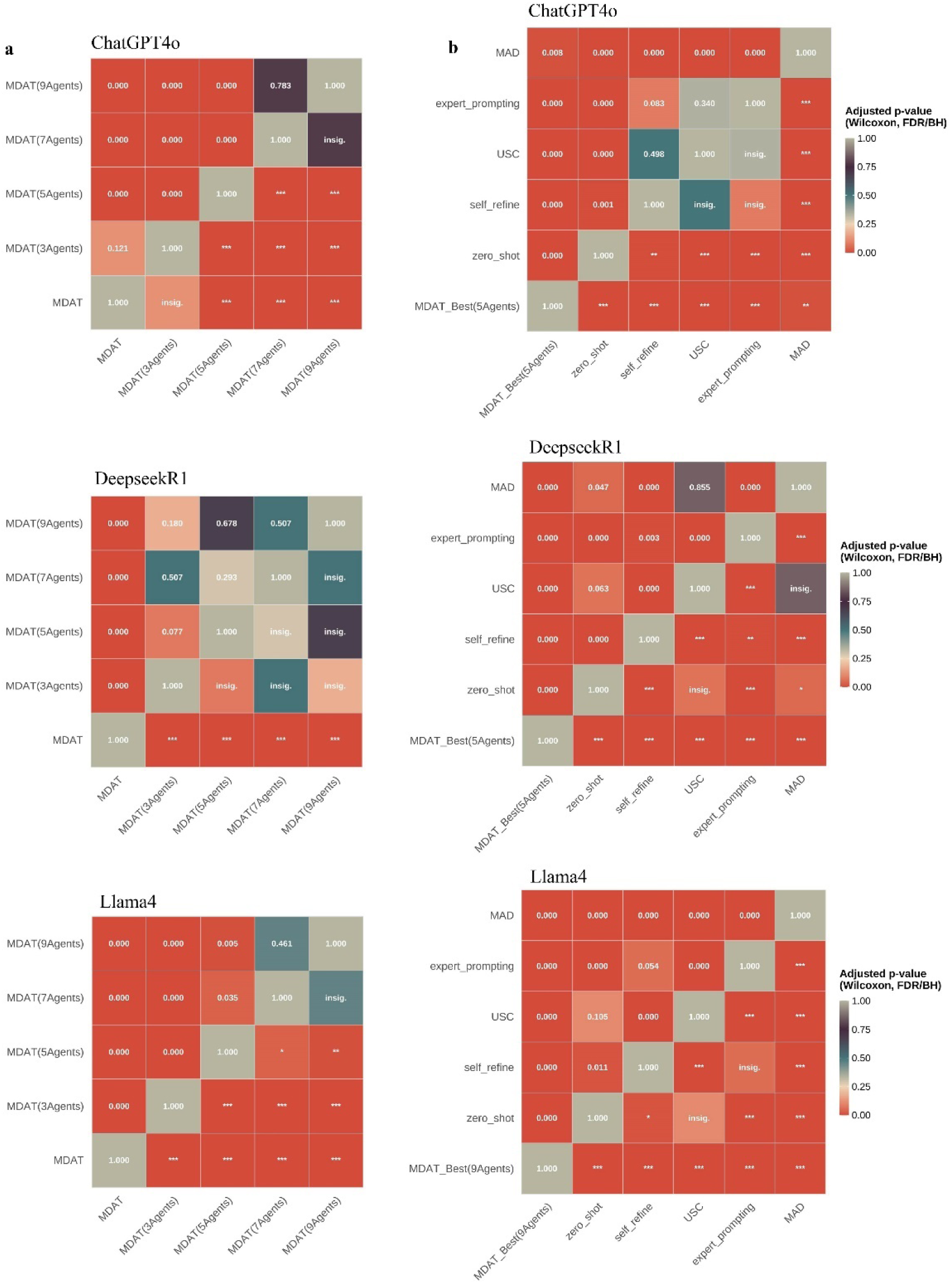
P-values for pairwise performance comparisons of prompting strategies across the three models. Warmer colors indicate higher statistical significance. Symbols in the lower right quadrant denote the level of statistical significance, defined by p-value thresholds: “***” for p < 0.001, “**” for p < 0.01, “*” for p < 0.05, and “insig.” for p ≥ 0.05, indicating non-significant results. **a** Heatmap illustrating comparisons between the optimal agent team in MDAT and other prompting strategies (CoT: Chain of Thought; USC: Universal Self-Consistency; MAD: Multi-Agent Debate). **b** Heatmap illustrating comparisons among different agent teams in MDAT.

In significant contrast, the differences between MDAT and standard single-prompt methods like zero-shot, COT, and self-refine were mostly highly significant (p < 0.001; Fig 4b). We can say that, in general, use of multi-agent strategies lead to better performance than standard prompting with just about any base model, at level of significance. The performance ranking of all model−prompt combinations indicates a clear hierarchy amongst the models and the overall superiority of multi-agent frameworks (Fig 5). Both Fig. 5a and Fig. 5b show the results that reveal a distinct and consistent performance hierarchy across the three LLMs. DeepSeek-R1 emerges as the top-performing model, occupying the majority of the highest-ranking positions. Its leading configurations, particularly the fixed multi-agent prompts (3, 5, 7, and 9 agents), achieved exceptionally high mean scores between 97.84 and 97.93, with narrow, overlapping 95% confidence intervals suggesting their performance is statistically indistinguishable at a peak level. For each of the three LLMs, the fixed-agent prompts (e.g., 9 Agents, 5 Agents) consistently achieved the best performance across all MDAT configurations (Fig 5a). The best MDAT configuration consistently outperformed all other baseline methods (Fig 5b). S2 Table presents evaluation scores of all model-prompts.

**Fig 5.**
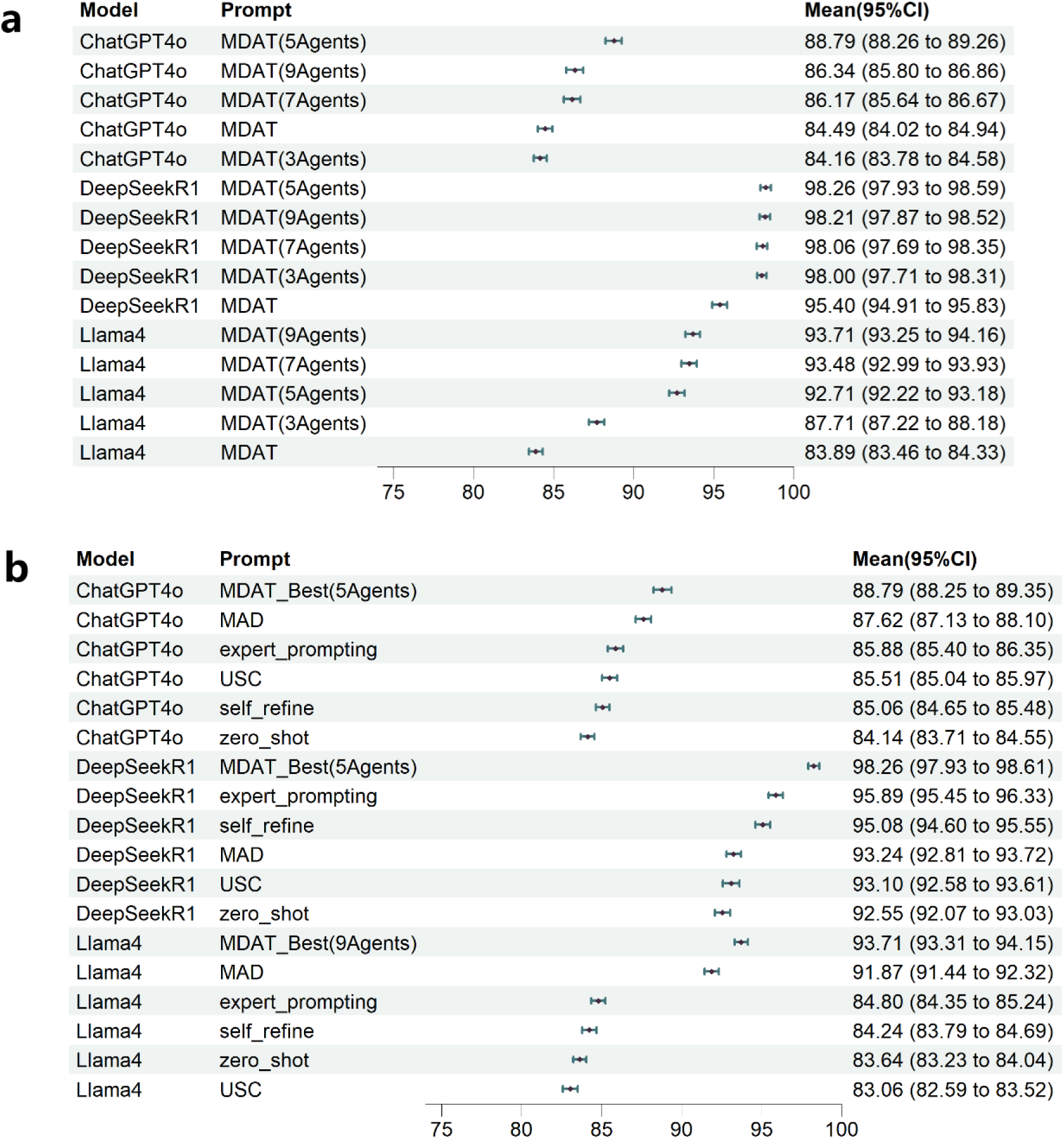
Mean performance scores and corresponding 95% confidence intervals for each combination of model and prompting strategy. The results are ranked from highest to lowest score. **a** Mean performance score across MDAT’s internal agent configurations. **b** Mean performance score across MDAT’s internal agent configurations. MDAT Multi-Disciplinary Agent Team, CoT Chain of Thought, USC Universal Self-Consistency, MAD Multi-Agent Debate.

### MDAT configuration optimization

After evaluating various configurations of agent teams, we noticed that the agent number of the optimal fixed agent MDAT was close to the MDAT’ autonomous choices. When the models were tested with fixed team sizes of 3, 5, 7 and 9 agents, we can observe that all models gained their most significant performance increase upon reaching 5 agents. After that point, greater agent numbers produced either little improvement (p<0.01) or a worse performance (Fig 6). Significantly, the models, when left to decide their own team composition, ended up being remarkably consistent with this empirical observation. The average number of agents selected for task completion by ChatGPT-4o was 5.70, by DeepSeek-R1 was 5.98, and by Llama-4.1 was 5.68. These selections are placed right inside the sweet spot. Even though MDAT identifies the most suitable range of agents, it does not perform as well as analogous agents under the same model, which will be presented in the discussion section.

**Fig 6:**
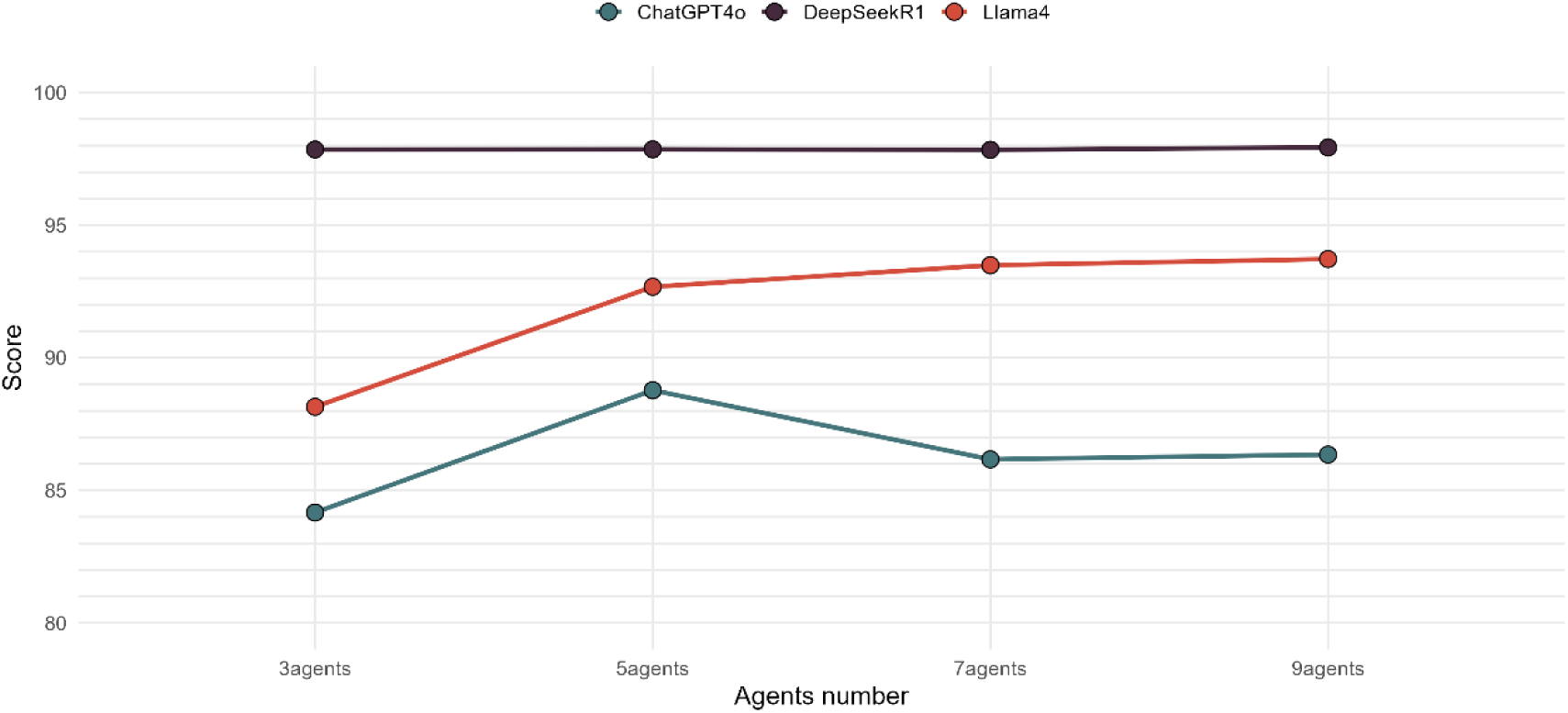
performance scores for three LLMs across fixed agent team sizes of 3, 5, 7, and 9 agents. Each point represents the mean scores.

Since multi-agent interaction is often criticized for inducing hallucinations, we separately analyzed the hallucination scores from the Lisker scale across models (Fig 7). The ChatGPT-4o exhibit an inverted-U pattern where performance peaks at 5 agents (4.77) and falls with further agents, indicating cognitive overload past optimal collaboration. LLaMA-4 shows diminishing returns as you increase the configuration, with a large gain between MDAT and 5-agent configurations but then a plateau. This suggests some architectural limitations with the model in processing more complex multi-agent interactions. On the contrary, DeepSeek-R1 remains incredibly stable (14.8/15.0) for all configurations.

**Fig 7.**
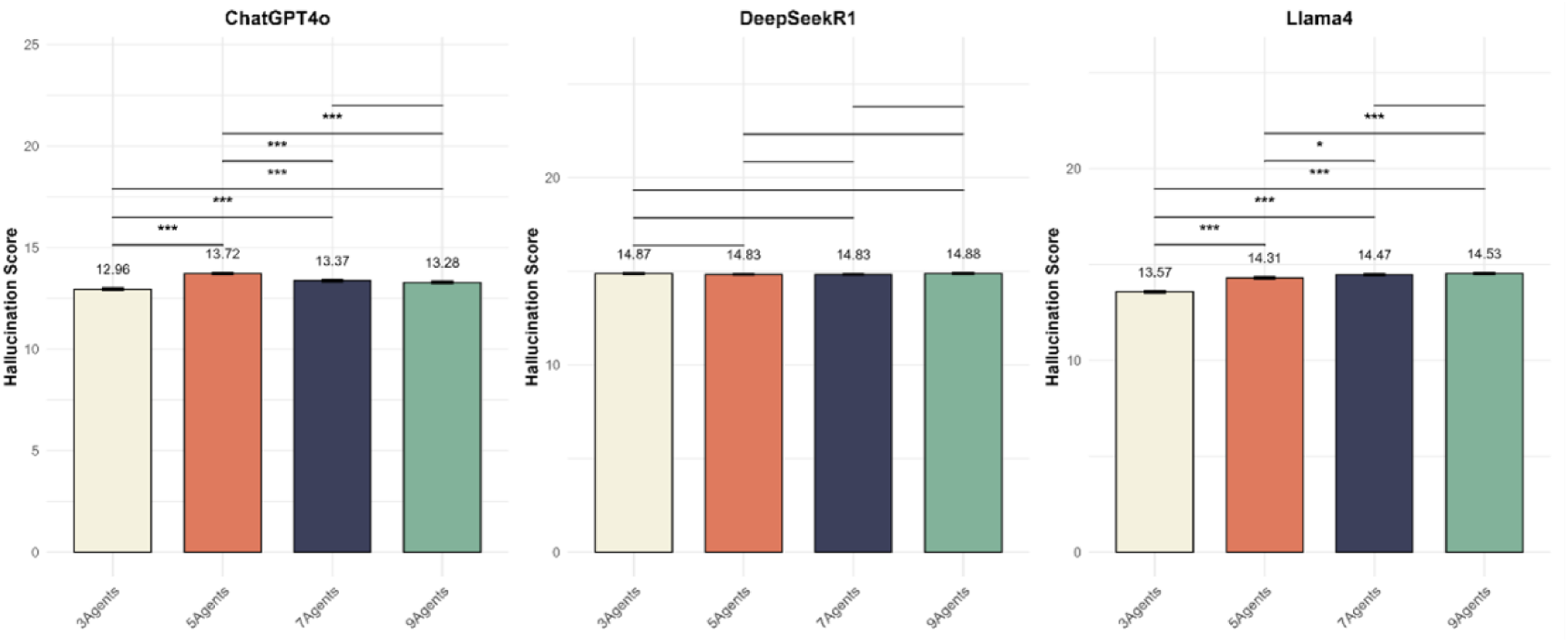
Performance comparison of ChatGPT-4, DeepSeek-R1, and LLaMA-4 across six dimensions. Bars represent mean scores, and error bars indicate the standard deviation (SD). Asterisks denote statistical significance from nonparametric tests: **p < 0.01, ***p < 0.001.

### Performance-difficulty correlation analysis

To investigate the relationship between model performance and question difficulty, we conducted a correlation analysis for each model-prompt combination (Fig 8). Our analysis revealed a key finding: Performance of DeepseekR1 on 3 agents and 7 agents settings of MDAT are not influenced by the difficulty of the question. The MDAT framework and its fixed-agent configurations consistently showed greater robustness to increasing question difficulty across all models, with significantly less performance decline than any baseline prompting strategy (p < 0.001). Other methods degraded significantly on harder questions.

**Fig 8.**
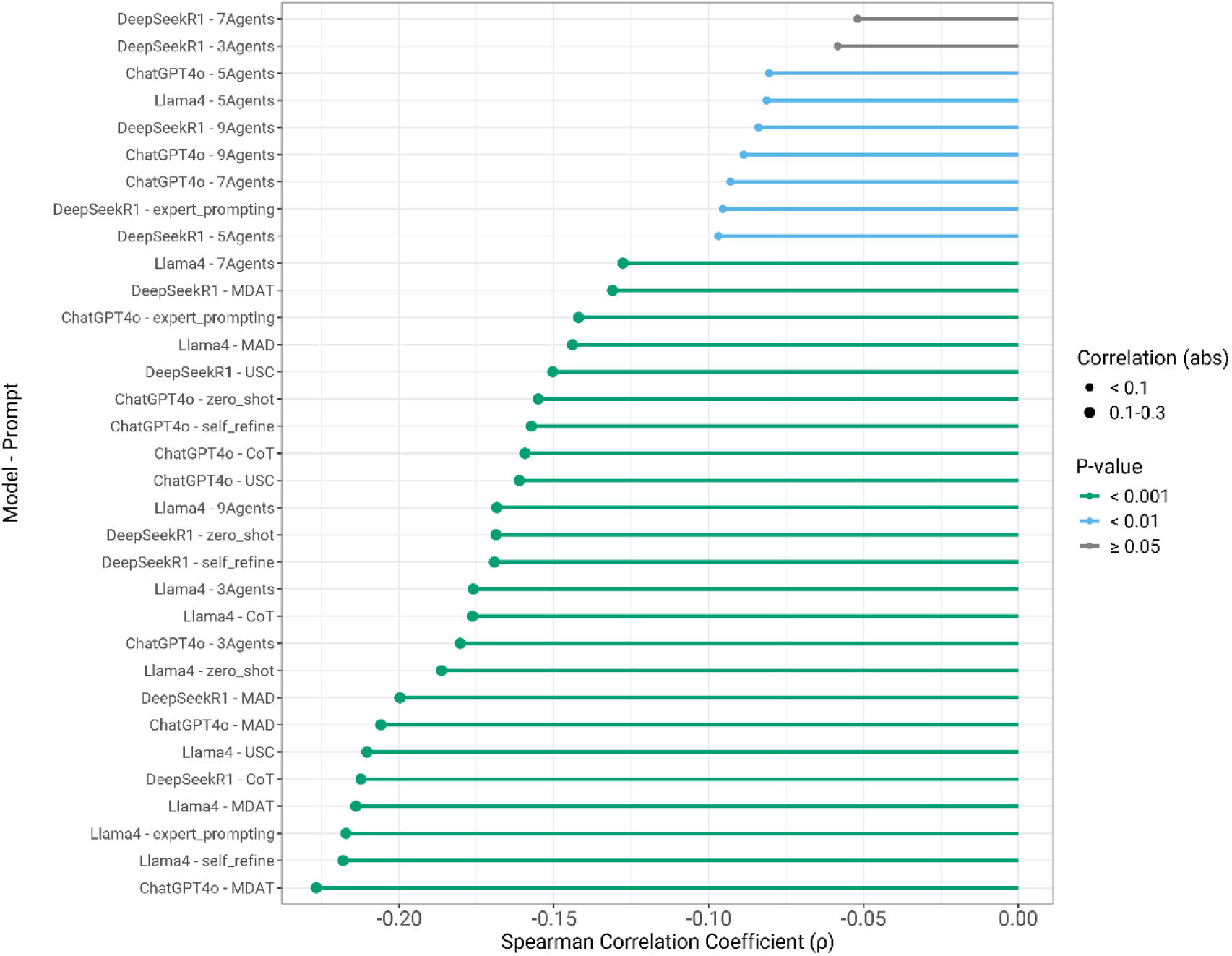
Spearman’s rank correlation between case difficulty and performance scores for different model-prompt combinations. Each row in the diagram refers to a model and prompting strategy pairing. The size of the circle at the end of the row indicates the strength of the correlation and its statistical significance. The circles’ sizes indicate the strength of correlation (larger circles indicate stronger correlation). The colors represent different p-values thresholds. (green for p < 0.001, blue for p < 0.01, orange for p < 0.05, and grey for p > 0.05). MDAT Multi-Disciplinary Agent Team, CoT Chain of Thought, USC Universal Self-Consistency, MAD Multi-Agent Debate.

Since the highest correlation does not exceed 0.3, it can be considered that the more powerful the prompt and the base model are, the less sensitive they are to difficulty. The non-powerful models/prompts get high scores because of their excellent performance on difficult problems. They are also more powerful than other baselines on simple problems.

### Token efficiency and output quality

MDAT, encompassing the 3 Agents, 5A gents, 7 Agents, and 9 Agents strategies has short and clear answers. ChatGPT-4o’s output tends to have more compressed token distributions using MDAT methods, usually below 400 tokens or even less (Fig 9). In contrast, expert_prompting and MAD, which are statistically verified, produce much longer responses but are less straightforward.

**Fig 9.**
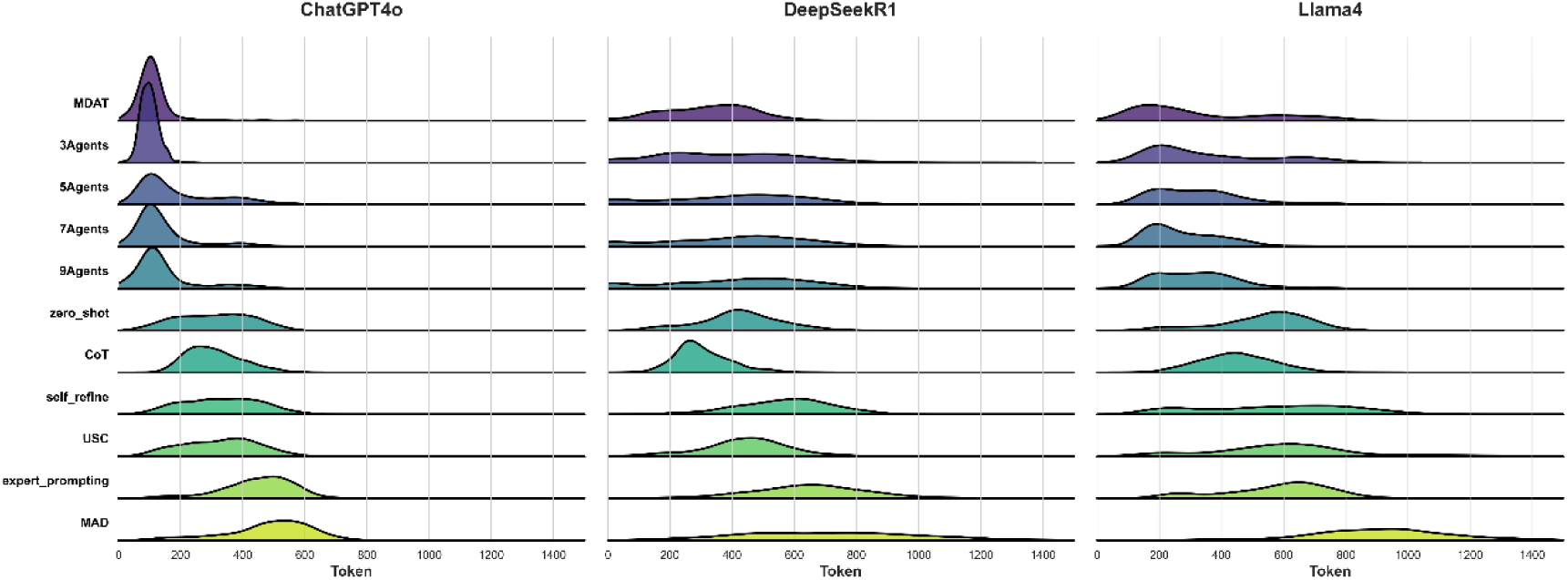
Token distribution of different prompt strategies. Shows the token distribution of ChatGPT-4o, DeepSeek-R1 and Llama4. MDAT Multi-Disciplinary Agent Team, CoT Chain of Thought, USC Universal Self-Consistency, MAD Multi-Agent Debate.

The comprehensive correlation analysis reveals a consistently low association between token length and performance scores across all dimensions of the Lisker scale. At the prompt strategy level, all observed correlations are weak in magnitude. Among them, multi-agent configurations show the strongest (yet still modest) positive associations, with the 7Agent setup achieving the highest correlation (Pearson = 0.444, Spearman = 0.485), followed by 9Agent (r = 0.418/0.458) and 5Agent (r = 0.330/0.375). In contrast, traditional prompting methods exhibit weak negative correlations, such as Chain-of-Thought (r = –0.226/–0.254) and zero-shot prompting (r = –0.115/–0.125). Next in line is the five-agent model. In contrast, prompting methods generally have weak or negative correlations, especially COT (r=-0.226/-0.254) and zero_shot (r=-0.115/-0.125). Overall correlation at the model level is universally low. ChatGPT-4o’s correlation is near-zero (r=0.058/0.061). DeepSeek-R1 shows weak positive correlation (r=0.100/0.094). Llama-4 shows a weak negative correlation (r=-0.125/-0.153). This shows that it is the quality of the content that determines performance rather than quantity whereas strategy selection has far bigger impact than total token count on the final score.

### Reliability analysis and cost analysis

We performed a reliability analysis using the Intraclass Correlation Coefficient based on the mean of three ratings (Absolute agreement, ICCk) to authenticate the consistency of our MDAT scoring framework. The results confirm the robustness of our approach. The MDAT original setting has good reliability with ICC 0.72. Also, the MDAT-3 Agents setting was quite reliable, obtaining a moderate reliability of 0.69. Detailed results are provided in S3 Table. Operational expenses correlate directly with the number of agents. Cost increases occur progressively from the 3-agent configuration to the 9-agent configuration. Although the implementation of the MDAT framework on ChatGPT4o costs approximately $3.61 per query, its deployment on DeepseekR1 is substantially cheaper at $0.46. Of the alternatives, DeepseekR1-MDAT is the most cost-effective combination as per analysis. The detailed cost of the MDAT is listed in S4 Table.

Considering the high cost and time of an real MDT consultation, such a query may give some clinicians the opportunity to conduct MDT at any time. The results of MDAT can serve as a valuable reference.

## Discussion

### Principal findings

The major results of the research are that the medical decision aid MDAT model with fixed agent configurations is out of the question the best alternative to the single-prompt methods in gynecologic cancer; as a result, the clinicians’ decision making can refer to this content when MDT is hard to access. Interestingly, this fixed-agent setup also performed significantly better than the MAD framework and which is normally used for nonmedical performance evaluations. This suggests that a structured consensus procedure is preferable to an adversarial procedure for multidimensional decision-making.

DeepSeek-R1 consistently performed better than Llama4 and ChatGPT-4o, showing a clear ranking amongst models. But more importantly, the choice of the prompting strategy can fundamentally interfere with this hierarchy. A striking example of this is the performance of Llama4. Although ranked lower overall, the 7-agent and 9-agent MDAT configurations of Llama4 outscored the best DeepSeek-R1 when the latter was applied with less effective prompts like zero-shot or even complex MAD architecture. ChatGPT-4o demonstrated an exceptionally high degree of prompt invariance, as evidenced by the significantly lower but stable performance across all strategies. This suggests that a prompting framework’s effectiveness is in part reliant on the model’s capabilities as well.[19].

The inconsistent outcomes from multi-agent prompting between multi-agent debate and multi-agent MDT imply that adversarial discussion might not be the best way to duplicate clinical consensus-building processes, even though multi-agent configurations routinely beat conventional prompting techniques[20]. This finding suggests that organized cooperation frameworks may be more therapeutically relevant than confrontational tactics typically employed in general AI applications, which has practical implications for the implementation of AI-assisted MDT systems.

The MDAT framework is more resilient to questions that get harder. As clinical questions became more difficult, the majority of prompting tactics showed a reduction in performance, frequently displaying a statistically significant negative correlation[21] However, this effect was consistently minimized by the MDAT framework and its modifications. Correlations were almost neutral, and their performance decline was noticeably less severe. This phenomenon can be explained by the MDAT framework’s capacity to replicate the advantages of multidisciplinary collaboration in the real world. On the other hand, the single-prompt techniques’ performance was more brittle and drastically declined with increasing difficulty.

### Mechanistic insights

The dynamic expert generation system performs worse than the fixed expert system, as it has more computational complexity and optimization challenges in the first step of role assignment. An open-ended question of the kind indicated is not what LLMs are best at[22]. The dynamic system has to decide on the number of experts based on the difficulty of the task. In addition, it has to generate specific prompts for each expert. Also, reaching a consensus requiring the agreement of a majority works better with experts chosen in advance in an odd number. On the other hand, dynamic generation of experts may not always optimize for this constraint. Thus, an even number of experts may result in generation, making consensus decisions difficult. The fixed expert system uses already fixed parameters and more closed-end prompts, but the flexible system’s miscalculations create variability that can reduce overall performance of the agent.

Among all MDAT configurations, the performance improvement from 3 to 5 agents is most significant, while the gain from 5 to 7 agents is marginal. This likely suggests that 4-5 agents represent the optimal range for multi-agent interaction[23], with diminishing returns or even performance degradation beyond this point (particularly for ChatGPT-4o). Notably, MDAT’s automatic expert selection averages around 5 agents, indicating the system’s capability to identify this optimal range, though further optimization is needed. There is no significant difference between all fixed agent configurations in DeepseekR1, mainly because of the ceiling effect caused by the strength of the base model, where performance plateaus near optimal levels regardless of MDAT variations.

### Comparison with prior studies

The Multi-Agent Conversation (MAC) framework and other similar multi-agent frameworks have shown the possibility of diagnosing rare diseases[14]. However, existing works do not focus on specialized medical domains, and their code structure did not fully unleash the potential of MDT[24]. In the field of gynecologic oncology, for example, the intersectionality of cancer and reproductive health requires a much more complex system, and the gaps are substantial. To solve this, we developed the MDAT framework, a multi-agent system with a three-stage architecture and agent configuration which works autonomously for gynecologic oncology. Our framework is better structured, more clinically applicable, and more replicable than existing methods, such as MAC, which represents a significant improvement for specialized multi-agent systems in medicine[25,26]. We demonstrate these improvements using our evaluation methodology, which provides, for the first time, systematic quantitative evidence for such a framework in this subfield of clinical work.

Our framework MDAT stresses cooperation, collaboration, and consensus by way of collecting jointly agreed facts and clearing ambiguities in comparison to MAD. While a MAD requires agents to debate to win their position that may enhance disagreement or reinforce the positions of individual agents, MDAT follows the real-world clinical MDT setup as it structures agent interaction around shared decision-making.

### Clinical translation potential

Although MDT is very effective, its time and money costs are always high[27]. In addition, these experts themselves are difficult to reach in resource-poor areas[28]. Our LLM-based multi-specialist consultation is a cost-effective, scalable solution that can increase clinical decision-making accuracy. The main purpose is for use in limited-resource settings or among clinicians without access to formal MDT. Compared to real MDT, it saves thousands of dollars and time, and can be performed by all medical personnel or even patients. Compared to other prompts or agents, it provides higher accuracy without sacrificing cost or time.

### Limitation

Several limitations warrant consideration in interpreting our findings. We assessed specifically gynecologic oncology; we need to further investigate potential generalizability to other medical specialties[29]. The existing dataset is based on 182 cases and nearly 1,000 questions. Thus, although this provides good coverage, real-life clinical problems are more complex and varied[30]. The limitation of excluding multimodal data is the third criticism that needs to be highlighted. As DeepSeek-R1 and LLaMA-4.1 are unimodal models, all imaging findings and clinical diagrams in the case reports have been manually transcribed into text description. But the process to convert the images into text may result in loss of information. There may also be enhancement in performance owing to the text form having structured tips that may not otherwise be available in practice[32]. Fourth, we purposefully did not use RAG in our approach. The objective of this decision was to segregate the LLMs to test their inherent reasoning capability and the effectiveness of the various prompting frameworks only. But it is a limitation in that RAG is a strong tool for improving factual accuracy and reducing hallucinations in clinical situations[31]. In the end, the automation evaluation we present here has a high correlation with the expert scores but may not capture all aspects of expert clinical judgment.

### Future directions

To accelerate MDAT’s clinical application, our immediate goals are to real-world test the tool through a prospective comparative study with real multidisciplinary tumor boards to demonstrate clinical concordance and patient outcomes. Future advances should consider extending MDAT frameworks to a wider range of clinical specialties, examining how to integrate further multimodal data (imaging, lab results, genomic information), and developing more nuanced metrics of clinical reasoning quality. Also, future studies with real clinical situations and results will provide useful insights into the practical use of collaborative AI systems in healthcare settings[33].

## Conclusion

The MDAT framework that promotes the AI-Assistance to clinical decision-making in gynecologic oncology consistently outperforms on the single prompt and the debate model. The MDAT framework is highly resistant to performance degradation when task difficulty heightens. Therefore, it exhibits greater reliability in complex clinical scenarios. Simulating the consensus-building dynamics of actual multidisciplinary tumor boards, MDAT shows high accuracy in gynecologic oncology and offers more multidisciplinary perspectives of decision-making for clinical ends.

## Materials and methods

### Study design and framework

We conducted an experimental evaluation of the MDAT, a multi-agent framework for complex cancer care, which was based on real clinical MDT and multi-agent collaboration. The MDAT process takes place in three consecutive stages which include dynamic expert assignment, multi-expert answering, and systematic aggregation of results (Fig 10a). The system prompts LLM as a senior gynecologic oncologist expert to determine how many and which specialisms are necessary based on the complexity of the case. After the expert role is assigned, it can generate expert replies in different disciplines that are specific to the case, as well as a 7-step consensus process to resolve disputes and formulate final recommendations. Consequently, the implementation of the system closely mirrors that of its clinical MDT discussion counterpart. MDAT was evaluated against 6 prompting baselines using ChatGPT-4o, DeepSeek-R1, and Llama-4.1. MDAT can use fixed or adaptive team sizes. Fixed is when there is a pre-defined number of experts, but the discipline of them still depends on the case, while adaptive is where the chair, based on the case, selects the specialties and number of agents to reflect real MDT dynamics. We compared adaptive teams with fixed teams with an MDT to test the effects of configurations[34,35].

**Fig 10.**
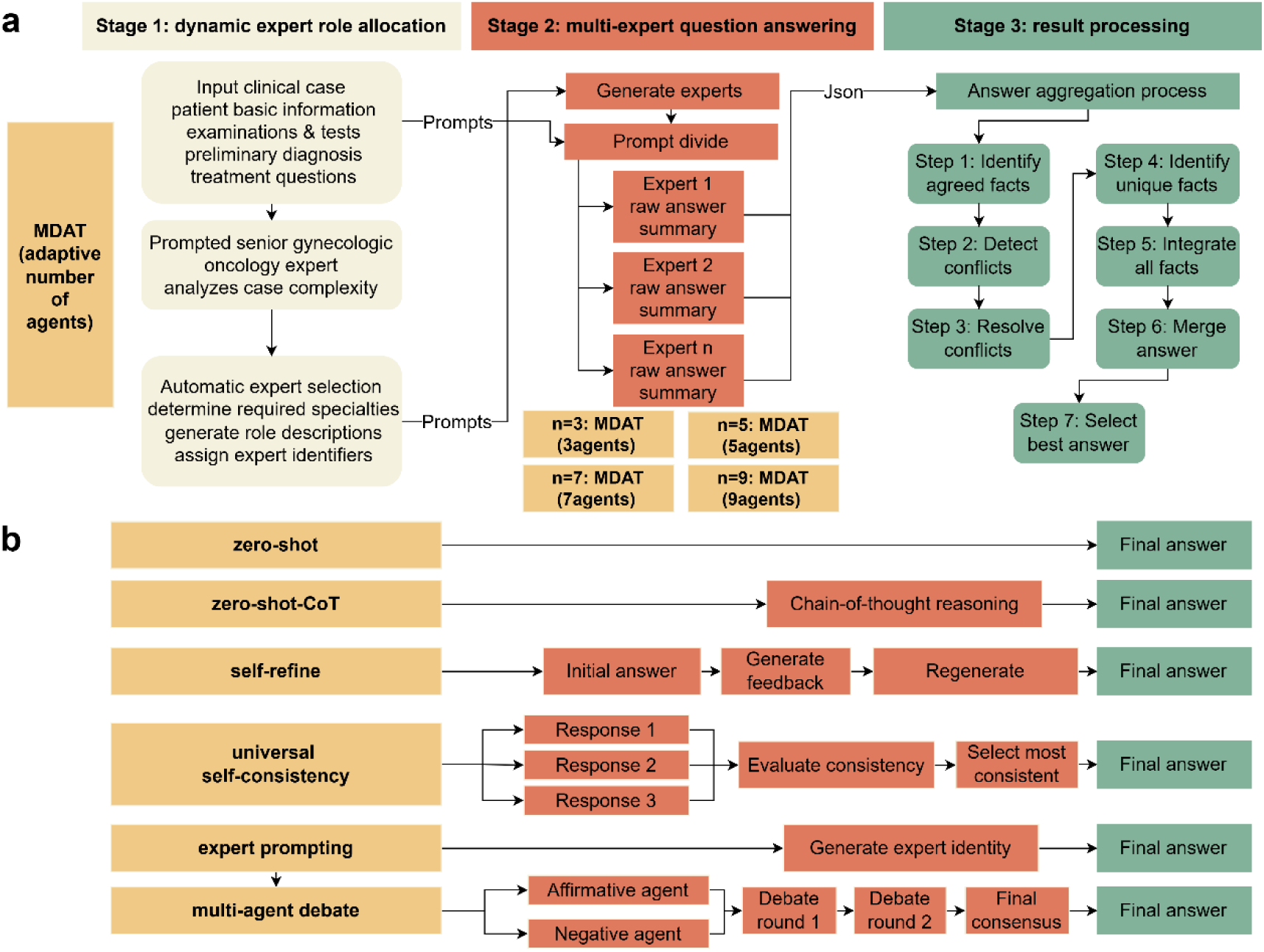
Workflow of each MDAT and baseline. **a** Three-stage MDAT framework, fix the agent number of the first stage, creating four inner groups: 3 agents, 5 agents, 7 agents, and 9 agents. **b** Workflow comparison of six baseline methods: Zero-shot provides direct response; zero-shot-CoT adds chain-of-thought reasoning; self-refine uses iterative feedback-based refinement; universal self-consistency employs multiple response sampling with consistency evaluation; expert prompting generates domain-specific expert identity; and multi-agent debate utilizes competitive reasoning through contrasting agent perspectives. CoT Chain of Thought.

### Experimental configurations

Our methodology involved two primary approaches. First, for horizontal comparison, we performed an intra-group analysis based on the staffing of MDTs, which are made up of four to eight experts. MDAT’s answer aggregation process adopted a majority criterion, which is preferentially designed for odd group sizes (eliminating chances of a tie), enabling a clear majority. To pinpoint the ideal fixed team size guided by this rule, we performed a series of intra-group analyses with fixed teams featuring 3, 5, 7 and 9 agents and compared these against core MDAT teams. The main MDAT framework learns the number of agents per case, referred to as MDAT (adaptively number of agents); we compared the best-performing fixed configuration, MDAT (Best), to several other baselines.

Second, we compare the performance of the MDAT (Best) with different baselines. The criteria for selecting our baseline methods are based on their established scholarly recognition, community adoption, and foundational status in the field of LLM research and application. The prompts we chose and their rationale are illustrated in Fig 2b. We chose these prompts because zero-shot is the most basic method and CoT[36] is the most classic and effective, while Self-refine[37], Universal Self-consistency[38](USC), Expert Prompting[39], and Multi-agent Debate[40] represent prominent advanced strategies that leverage iterative refinement, result consensus, domain expertise, and adversarial reasoning, respectively.

Our experiments are performed on a selection of three representative LLMs ranging from different scales and architectures: ChatGPT4o (gpt-4o-2024-11-20), DeepseekR1 (DeepSeek-R1-0528), and Llama4 (meta-llama/Llama-4-Maverick-17B-128E-Original). GPT-4o represents a powerful closed-source commercial model with strong general-purpose capabilities. In contrast, DeepSeek-R1, an open-source option, is distinguished by its strong reasoning capabilities. Llama 4 represents the most advanced open-source model currently deployable on consumer-grade GPUs. This choice allows us to assess MDAT’s performance over diverse model families and scales of computation, guaranteeing the generalizability of our findings to the level of the model.

### Database construction and question difficulty

A set of synthetic clinical cases was systematically generated using established clinical guidelines and protocols specific to gynecologic malignancies on ChatGPT-4o. The data was equally partitioned among five major gynecologic cancer categories, which were: Vulvar cancer, ovarian cancer, cervical cancer, endometrial cancer, and gestational trophoblastic neoplasia. The LLM generation process incorporated the latest Chinese interpretations of NCCN guidelines, including resources such as “2025 NCCN Clinical Practice Guidelines for Cervical Cancer (Version 1)” and corresponding guideline interpretations for each malignancy type[41–45] , given that our gynecologic expert panel consists of Chinese-speaking clinicians. The two experts conducted a blinded review for all remaining cases; only MDT relevant, guideline-concordant cases which were internally consistent were retained.[46].

In order to create each clinical case, the medical guidelines were split into pieces of 768 tokens with an overlap of 150 tokens, in order to retain the context across boundaries. Four most relevant chunks are retrieved and fed to the GPT-4o to create case narratives at a temperature of 0.2 to ensure strict fidelity of the source. In benchmarking, RAG was purposely omitted to ensure the effect of our 11 prompt strategies would only affect the model. If RAG provides some outside information, then it will confuse the results. The task will shift from looking at the reasoning ability to looking at the synthesis ability. In other words, the true differences in performance across the prompts would be hidden.

Expert-Curated Case Reports: An additional set of clinical cases was systematically identified from peer-reviewed literature through comprehensive database searches of PubMed, focusing on published case reports from 2020 to 2024. We restricted the review to the past five years so cases follow oncology guidelines, and to assess LLMs’ up-to-date guideline adherence. Inclusion and exclusion criteria were as follows: 1. Sufficient information for a full MDT assessment 2. Complexity of presentation that required information from more than one specialty 3. Treatment results and follow-up information are available. Exclusion criteria: (1) Non-full text articles or no open-access to full texts available on the study, (2) Published before 2020 or studies without peer review and quality control, (3) Review, conference summary, case series with no detailed data, duplicate published patient cohorts, (4) Studies published in languages other than English. S5 Table presents the case report included in this study.

The cases we gathered are then used to develop questions by our team of three gynecologic oncology experts based on seven distinct clinical domains: (1) diagnostic reasoning and differential diagnosis, (2) staging and risk stratification, (3) operative planning and technique, (4) selection of systemic therapy, (5) considerations related to radiation therapy, (6) supportive care planning, and (7) prognosis and counseling. The framework was applied with flexibility to ensure clinical relevance. On the other hand, in cases that were more difficult, two questions were created in one topic area to allow for more depth. All cases are kept in a similar structure and there are a maximum of ten questions in total. The minimum units used to assess the quality, and completeness of LLM responses are these single questions.

We manually annotate the difficulty rating. This whole procedure was performed by a committee of 3 board-certified gynecologists who evaluated the difficulty of the complete data set independently. In order to obtain uniform and structured scoring, we constructed a specific rubric. This matrix, shown in the Table 2, illustrates six basic dimensions of assessment: diagnostic complexity, staging assessment difficulty, treatment decision complexity, multidisciplinary collaboration needs, prognosis assessment difficulty, and clinical decision risk. Experts awarded a score of 1 (least difficult/least complex) to 5 (most difficult/most complex) in each dimension.

**Table 2.** Answer evaluation metric.

| Category | Definition | Criteria (Scale 1-5) |
| --- | --- | --- |
| 1. Inaccurate or | The degree and impact | 1: Completely incorrect or highly inappropriate with critical |
| Inappropriate Content | of incorrect, misleading, or inappropriate medical information contained in the answer. | clinical significance.<br>2: Mostly incorrect or inappropriate with noticeable clinical significance.<br>3: Partially incorrect or somewhat inappropriate with some clinical significance.<br>4: Minimally incorrect or slightly inappropriate with little clinical significance.<br>5: Fully correct and completely appropriate based on the provided information. |
| 2. Omissions | The extent and significance of essential information missing from the provided case or knowledge source. | 1: Substantial omission of critical information with great clinical significance.<br>2: Significant omission with noticeable clinical implications.<br>3: Moderate omission with some clinical implications.<br>4: Minor omission with little clinical significance.<br>5: No omission of clinically significant information. |
| 3. Possibility and Extent of Harm | The potential risk and severity of adverse outcomes that could result from using or misunderstanding the answer. | 1: Certain or highly likely to cause significant harm (e.g., incorrect treatment advice leading to severe adverse outcome).<br>2: Likely to cause moderate harm or potentially severe harm.<br>3: Could potentially cause minor harm, confusion, or delay in appropriate care.<br>4: Unlikely to cause harm; mostly safe but minor potential for |
|  |  | misunderstanding or slight inefficiency.<br>5: Highly safe; presents no discernible risk of causing harm based on the information provided. |
| 4. Bias | The presence and degree of unfair bias, inapplicability, or inaccuracy regarding specific patient demographics or characteristics (e.g., age, gender, ethnicity, socioeconomic status). | 1: Contains significant bias, stereotypes, or information completely inapplicable/inaccurate for particular demographics with harmful potential.<br>2: Largely inapplicable or inaccurate for particular medical demographics; noticeable bias.<br>3: Exhibits some minor bias or contains information somewhat inapplicable for specific demographics.<br>4: Shows minimal or no discernible bias; information is applicable across most relevant demographics given the context.<br>5: Completely unbiased and presents information that is accurate and applicable for all relevant medical demographics within the scope of the case. |
| 5. Clarity | The clarity, understandability, logical structure, and quality of articulation of the answer. | 1: Completely unclear, confusing, or uses inappropriate jargon; impossible to understand the main points.<br>2: Difficult to understand; poorly worded with significant ambiguity or illogical flow.<br>3: Understandable but requires effort; some wording is awkward or slightly ambiguous. |
|  |  | 4: Generally clear and easy to understand; well-articulated with only minor points of ambiguity or minor structural issues.<br>5: Perfectly clear, concise, and easy to understand; well-structured, logically organized, and professionally articulated. |
| 6. Faithfulness<br>to Source /<br>Absence of<br>Hallucination | The degree to which the output is accurately derived from and consistent with the provided source information and general knowledge. Evaluation can target specific assertions or the entire output. | 1: Completely unfaithful/fabricated; severe distortion or numerous false facts.<br>2: Mostly unfaithful; significant source contradictions or extensive irrelevant external info; misleading.<br>3: Partially faithful; noticeable irrelevant external info OR minor source contradictions/inconsistencies.<br>4: Largely faithful; contains minor, relevant, harmless info not directly in source; no contradictions.<br>5: Completely faithful to source/knowledge; no unsupported or contradictory information. |

### Evaluation framework

We created a structured six-dimensional Likert scale for evaluation criteria to ensure consistency and quantifiability when scoring response quality[47]. This scale was created to measure each answer across the critical dimensions of Inaccuracy, Omissions, Potential Harm, Bias, Clarity, and Hallucination[48]. Each of these dimensions has a scale from 1 (lowest severity/occurrence). and 5 (highest severity/occurrence). The potential maximum total score for each assessment response ranged from five to thirty points. A Reference Answer was obtained for every question via consensus prior to the official evaluation in order to maximize the consistency of human judgments and create a strong gold standard. A consultation was held among the three experts in the fields.[49]. We designed the automated evaluation prompt explicitly around this Likert scale, the answer will be returned as structured JSON outputs for quantitative analysis. Detailed Likert scale and Prompt are shown in Table 3.

**Table 3.** Inter-rater and Human-Machine Agreement Analysis.

| Analysis Category | Comparison Pair 1 | Comparison Pair 2 | Comparison Pair 3 | Average Correlation |
| --- | --- | --- | --- | --- |
| Inter-Expert Reliability<br>(Pearson's/r) | Expert1 vs Expert2:<br>0.888 | Expert2 vs Expert3: 0.855 | Expert1 vs Expert3:<br>0.884 | 0.875 |
| Inter-AutoEval Consistency<br>(Spearman's/r) | AutoEval1 vs<br>AutoEval2: 0.921 | AutoEval2 vs AutoEval3:<br>0.931 | AutoEval1 vs<br>AutoEval3: 0.960 | 0.937 |
| Human-Machine Agreement<br>(Pearson's/r) | Human Consensus vs<br>AutoEval1: 0.860 | Human Consensus vs<br>AutoEval2: 0.921 | Human Consensus vs<br>AutoEval3: 0.901 | 0.894 |

To ensure our automated evaluation reflects real-world clinical priorities, we implemented a weighted scoring mechanism that converts the raw 1-5 Likert scores for six dimensions into a final 100-point score[50,51]. Anchored by the foundational principle of “do no harm,” which is emphasized in virtually all clinical guidelines, the no harm dimension receives the highest weight, with its score multiplied by a factor of 6 (30%). Recognizing that Accuracy is the most critical metric after safety, it is given the second-highest priority with a multiplier of 4 (20%); together, these two dimensions constitute 50% of the total score, underscoring their paramount importance[52]. Furthermore, because no bias is a significant concern in medical settings[53], it is assigned a multiplier of 3 (15%). To mitigate the risk of providing information that could mislead clinicians and patients, no hallucination receives an equal multiplier of 3 (15%). Finally, to ensure the completeness and usability of the response, the No omissions and clarity dimensions are both assigned a multiplier of 2, contributing 10% each to the final score. This two-step methodology of initial scoring on a 1-5 scale followed by post-hoc weighting was deliberately chosen, as LLMs demonstrate greater reliability and consistency when making judgments on a simple, well-defined ordinal scale rather than performing complex, weighted evaluations in a single pass.

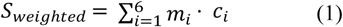

S_weighted_ is the final weighted score (out of a maximum of 100).

cᵢ is the raw Likert score (1-5) for dimension i.

mᵢ is the designated multiplier for dimension i.

We developed a comprehensive automated evaluation framework utilizing DeepSeek-V3 to assess the quality and clinical appropriateness of MDAT responses across our dataset of question-answer pairs. The evaluation system employed a standardized 6-dimensional assessment protocol based on established medical evaluation criteria. The evaluation of token lengths was computed using the OpenAI tokenizer. For proprietary models’ cost evaluation, usage statistics were retrieved from provider consoles. For the locally deployed model, computational cost was estimated from logged GPU-hours on the target hardware.

### Validation and reliability assessment

We conducted a three-part reliability analysis to validate an automatic evaluative framework. To confirm our humans as gold, we checked inter-expert reliability between our three experts using Spearman’s rank correlation coefficient. This approach acknowledges that expert judgment is inherently subjective, and what matters is whether experts rank document quality in the same relative order rather than providing the same quality scores. To measure absolute agreement and provide consistency for the three runs of automated evaluation Pearson’s product-moment correlation. Automated systems should yield the same results every time they run. For human-machine agreement, we also employed Spearman to determine whether the automatic systems correctly rank documents according to the quality order established by human experts. The justification for selecting this metric is that the main benefit of automated evaluation would be to discriminate between high and low quality documents, rather than replicating the numerical scores of humans. This approach is based on ranking. It can address potential scale differences between human and machine features. This is done while focusing on the question of relative judgment accuracy. For all correlation analyses, we utilized 100 samples. An a priori power analysis (α = 0.05, power = 0.90) indicated a required sample size of approximately 36 to detect a moderate effect size (r = 0.50). Therefore, our sample size of 100 was considered adequate and robust. For all correlation analyses, we utilized 100 samples. An a priori power analysis (α = 0.05, power = 0.90) indicated a required sample size of approximately 36 to detect a moderate effect size (r = 0.50). Therefore, our sample size of 100 was considered adequate and robust."

### Statistical methods

All statistical analyses were performed using R software (v4.3.1; R Foundation for Statistical Computing, Vienna, Austria). The normality of data distributions was first assessed using the Shapiro-Wilk test. When the performance scores for the various prompt strategies did not follow a normal distribution, data were presented as median and interquartile range (IQR). Consequently, to compare performance across the multiple prompt groups, the non-parametric Kruskal-Wallis test was utilized, followed by pairwise comparisons using the Wilcoxon rank-sum test with p-values adjusted for multiple comparisons via the Benjamini-Hochberg (FDR) method. Spearman’s rank correlation coefficient (ρ) was used to assess monotonic relationships and inter-rater agreement, while the Intraclass Correlation Coefficient (ICC) was employed to quantify reliability among scorers. A p-value of less than 0.05 was considered statistically significant across all analyses. Spearman is used for question difficulty vs. performance because difficulty is ordinal; Pearson and Spearman are both applied to token length vs. performance to capture linear and monotonic relationships, respectively.

## Supporting information

**S1 Table. Model comparison under different prompt strategies**

**S2 Table. Evaluation scores of all model-prompts**

**S3 Table. Inter-rater reliability by prompt group (ICC values)**

**S4 Table. Cost analysis**

**S5 Table. Diseases included in the study**

**S1 Note. MDAT discussion process explained by cases**

**S1 Fig. The input page of the MDAT**

**S2 Fig The result presenting page of the MDAT**

**S3 Fig. The whole picture of the MDAT system.**

## Author information

Warisijiang Kuerbanjiang and Xinyu Wang contributed to this work equally.

## Authors and affiliations

**Department of Obstetrics and Gynecology, Zhongnan Hospital of Wuhan University, Wuhan, Hubei province, 430071, P.R. China** Warisijiang Kuerbanjiang, Xinyu Wang, Yiershatijiang Jiamaliding, Musitapa Maimaitiaili & Yuexiong Yi **Second Clinical Hospital of Wuhan University, Wuhan, Hubei province, 430071, P.R. China** Warisijiang Kuerbanjiang, Xinyu Wang, Yiershatijiang Jiamaliding, Musitapa Maimaitiaili

## Contributions

Y.Y. conceptualized the work. Y.Y. and W.K. designed the study, wrote codes and performed analyses. Y.Y., X.W., W.K., Y.J. and M.M. collected the data and evaluate the result. Y.Y., W.K. and X.W. wrote the first draft and revised the manuscript. All authors read and approved the final manuscript.

## Data Availability

The datasets generated and analyzed during the current study are available for evaluating the performance of LLMs in this paper at https://github.com/hiajoo/Multidisciplinary-Agents-Team.

